# Sealing Behaviour in Transcatheter Bicuspid and Tricuspid Aortic Valves Replacement through Patient-Specific Computational Modeling

**DOI:** 10.1101/2021.05.07.21256799

**Authors:** Xian-Bao Liu, Jia-Qi Fan, Peter Mortier, Yu-Xin He, Qi-Feng Zhu, Yu-Chao Guo, Xin-Ping Lin, Hua-Jun Li, Ju-Bo Jiang, Giorgia Rocatello, Vanda Oliveira, Tim Dezutter, Lars Sondergaard, Jian-An Wang

**Author notes:** **Address for Correspondence:** Jian’an Wang, MD, PhD Corresponding author; E-mail address (J. Wang)., Mailing address: Department of Cardiology, Second Affiliated Hospital, Zhejiang University, School of Medicine, Hangzhou 310009, China. Drs Liu and Fan contributed equally to the manuscript.

## Abstract

**BACKGROUND:** Patient-specific computer simulation of transcatheter aortic valve replacement (TAVR) can provide unique insights in device-patient interaction. The aim of this study was to compare transcatheter aortic valve sealing behaviour in patients with bicuspid aortic valves (BAV) and tricuspid aortic valves (TAV) through patient-specific computational modeling.

**METHODS:** Patient-specific computer simulation was retrospectively performed with FEops HEARTguide for patients who underwent TAVR using the Venus A-Valve. Simulation output was compared with postprocedural computed tomography and echocardiography to validate the accuracy of the simulation. Sealing behaviour was analysed based on the predicted device-patient interaction by quantifying the distance between the transcatheter heart valve (THV) skirt and the surrounding anatomical regions. Skirt malapposition was defined by a distance larger than 1mm.

**RESULTS:** In total, 43 patients were included in the study. Predicted and observed THV frame deformation showed good correlation (R^2^ ≥ 0.90) for all analysed measurements (maximum diameter, minimum diameter, area and perimeter). The amount of predicted THV skirt malapposition was strongly linked with the echocardiographic grading of paravalvular leakage (PVL). More THV skirt malapposition was observed for BAV cases when compared to TAV cases (22.7% vs 15.5%, p < 0.05). A detailed analysis of skirt malapposition showed a higher degree of malapposition in the interleaflet triangles section for BAV cases as compared to TAV patients (11.1% vs 5.8%, p < 0.05).

**CONCLUSIONS:** Patient-specific computer simulation of TAVR can accurately predict the behaviour of the self-expanding Venus A-Valve. BAV patients are associated with more malapposition of the THV skirt as compared to TAV patients, and this is mainly driven by more malapposition in the interleaflet triangle region.

**Key messages:** *What is already known on this subject?:* Transcatheter aortic valve replacement (TAVR) in patients with bicuspid aortic valve (BAV) is becoming increasingly important due to expanding indications towards younger patients, as well as to geographic growth of the therapy. Patient-specific computational modeling of TAVR based on pre-procedural computed tomography (CT) has emerged as a promising technology capable of accurately predicting device-anatomy interaction, as well as paravalvular leakage and the risk on TAVR-induced conduction abnormalities for both tricuspid aortic valve (TAV) and BAV patients.

*What might this study add?:* BAV patients are associated with more malapposition of the transcatheter heart valve (THV) skirt as compared to TAV patients, and this is mainly driven by more malapposition in the interleaflet triangle region. More cases and studies are needed to confirm the results and related malapposition of the THV skirt to clinical echo-based paravalvular leakage grading.

*How might this impact on clinical practice?:* Patient-specific computational modeling of TAVR based on pre-procedural CT might be performed in BAV patients. Transcatheter heart valve size and ideal implanted depth could be recommended to reduce the malapposition and potential paravalvular leakage.

## Introduction

Transcatheter aortic valve replacement (TAVR) in patients with bicuspid aortic valve (BAV) is becoming increasingly important due to expanding indications which include younger patients, as well as to global adoption. In many countries, TAVR has become the standard of care for high risk patients, and is now expanding into younger, lower risk patients, resulting in an increased amount of patients with BAV stenosis [1–4]. On the other hand, the Chinese TAVR market is still relatively small but growing rapidly, and the prevalence of BAV cases in China is notably higher than in other countries [5]. Several clinical studies have demonstrated the safety and efficacy of TAVR in BAV patients [3,4], but there are still several challenges when treating BAV stenosis and patients should be carefully selected. Therefore, efforts to increase our knowledge of how TAVR devices interact with BAVs remain important.

The interaction of transcatheter heart valves with the aortic root is likely to be different between BAV and tricuspid aortic valve (TAV) patients. While device sizing for TAV cases is mainly based on the dimensions of the aortic annulus, an assessment of the supra-annular structure seems mandatory for BAV cases as this can be the primary location where the THV interacts with the aortic root [6,7]. This, however, depends on several anatomical factors such as BAV type, calcium burden, raphe length and the ratio of the intercommissural diameter to the mean annular diameter [7].

Patient-specific computational modeling of TAVR with FEops HEARTguide (FEops, Ghent, Belgium) based on pre-procedural computed tomography (CT) has emerged as a promising technology capable of accurately predicting device-anatomy interaction, as well as paravalvular leakage and the risk on TAVR-induced conduction abnormalities for both TAV and BAV patients [8–12]. Validation data is mainly available for the Medtronic self-expanding and the Boston Scientific mechanically expandable THVs. These three-dimensional computer models provide detailed insights that cannot be obtained through post-procedural imaging, and may also help to better understand the sealing behaviour in BAV and TAV patients.

In this study, we aimed to validate a patient-specific computer simulation of TAVR in Chinese patients treated with the self-expandable Venus-A valve and use the validated computational model to explore potential differences in the sealing behaviour between BAV and TAV patients.

## Methods

A retrospective single-centre study was performed on patients who underwent transcatheter aortic valve replacement using a Venus A-Valve (Venus Medtech). Both pre- and post-procedural CT imaging was available for all patients. All dual source computed tomography (DSCT) examinations were performed with the second generation dual-source CT (SOMATOM Definition Flash, Siemens Medical Solutions, Germany). The scan area was craniocaudal from the subclavian artery to the iliofemoral branches. Prospective ECG gating with a pitch of 2.4 was performed. Around 60–80 ml of iodine-containing contrast agent (Omnipaque 370 mg I/ml, GE Healthcare, Shanghai, China) was injected with a dual head power injector (Mallinckrodt, American) at a flow rate of 4 ml/s followed by 60 ml 0.9% saline solution at the same flow rate. A bolus tracking method was used in the descending aorta with a pre-set threshold of 180 Hounsfield Units (HU) to achieve optimal synchronization. The tube voltage was 100 kV, with a reference tube current-time product of 280 mAs and a collimation of 38.4 mm (2*32*0.6 mm3) with double sampling by z-axis flying focal spot. All procedures were performed as reported in previous studies [13,14]. The study was approved by the medical ethics committee of Second Affiliated Hospital of Zhejiang University and carried out according to the principles of the Declaration of Helsinki. All patients provided written informed consent for TAVR and the use of anonymous clinical, procedural, and follow-up data for research.

### Virtual Device Modeling

Accurate finite element models of the frames of all Venus A-Valve sizes (23, 26, 29 and 32mm) were generated based on CAD (Computer Aided Design) data provided by the device manufacturer. A virtual radial force test was performed to validate the virtual device models using the finite element analysis (FEA) software Abaqus (Abaqus v6.12, Dassault Systèmes, Paris, France). For this test, the device was crimped to a smaller diameter (loading) and then released (unloading) while the radial force in the crimper was measured. The model radial force was then compared with the experimental radial force data during unloading and within the relevant deployment range for each valve size. Model parameters were calibrated until excellent agreement was obtained.

### Patient-specific Computational Modeling

Three-dimensional patient-specific geometries of the native aortic root (including the calcified native leaflets) were reconstructed from pre-operative contrast-enhanced CT scans, using the image segmentation software Mimics (Mimics v21.0, Materialise, Leuven, Belgium). Venus A-valve models were then virtually deployed in these geometries using Abaqus. These simulations allow to assess the device, native leaflet and aortic wall deformation as previously described [8,10,11]. For each simulated implantation, the valve size selection and the depth of implantation were aligned with the clinical procedure. The simulated depth of implantation was iteratively adjusted to match the actual depth of implantation derived from the post-operative geometry, which was reconstructed from post-operative CT images using Mimics. This was done by overlaying the simulation results with the post-operative geometry using a manual geometrical registration method. An overview of these different reconstruction and modeling steps is summarized in Figure 1.

**Figure 1.**
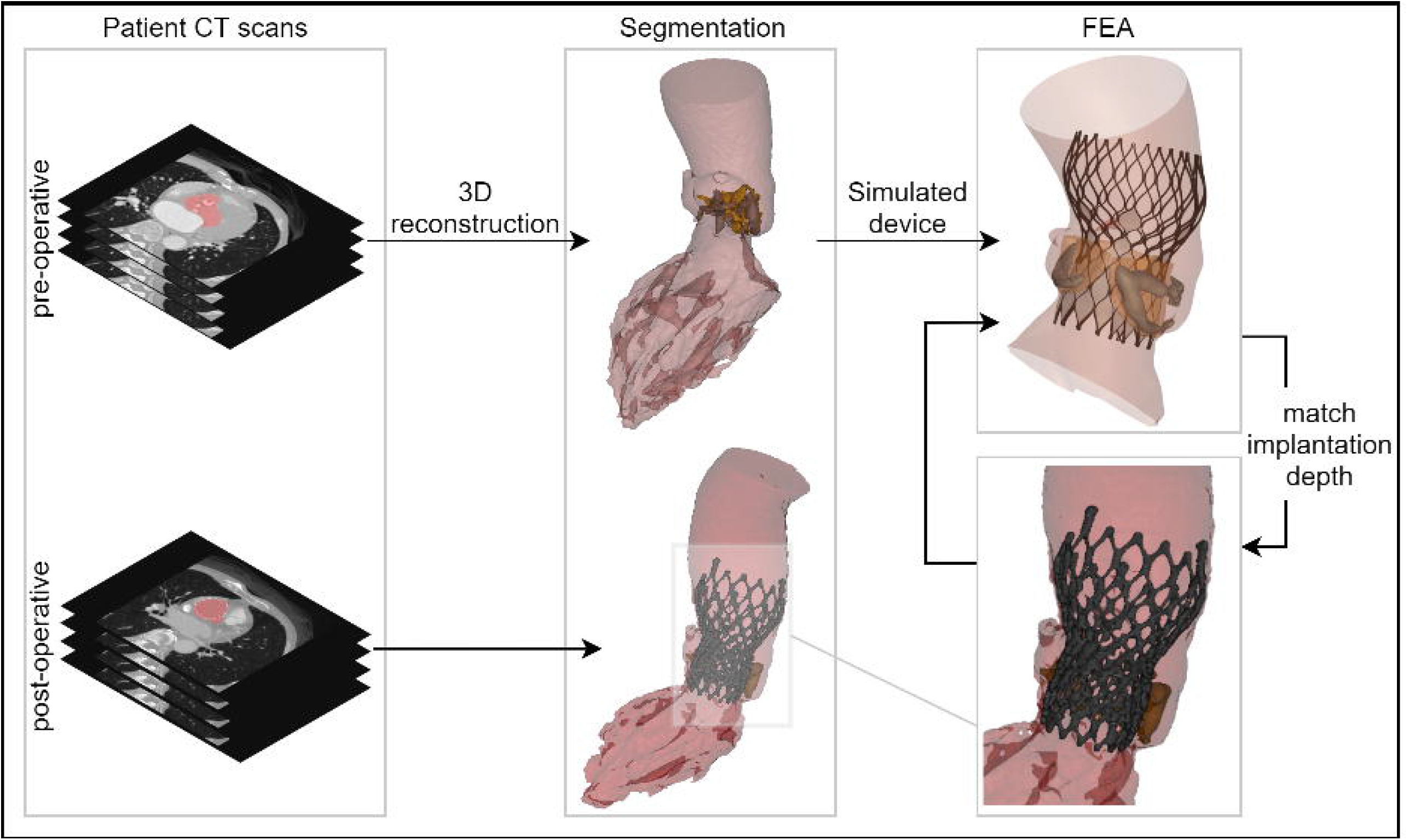
Overview showing the main steps of the virtual insertion of a Venus A-valve in a patient specific geometry derived from pre-operative CT, while aiming for a virtual implantation depth identical to the actual one. CT = computed tomography; 3D = three-dimensional; FEA = finite element analysis

### Frame deformation comparison

For each patient, predicted frame deformation was both qualitatively and quantitatively compared to the post-operative device deformation (CT). A visual inspection was performed by overlaying the predicted and post-operative devices and their dimensions (minimum and maximum diameter, perimeter and area) were quantified at four relevant device levels: commissures, central coaptation, nadir and ventricular end [8].

### Sealing analysis

The regions of skirt apposition and malapposition were determined for all patients using the predicted device and aortic root deformation. Apposition was considered when the deformed device skirt is in contact with the anatomy, while malapposition when the opposite is verified. The areas corresponding to the apposed and malapposed skirt were quantified in four different regions of the aortic root anatomy: left ventricular outflow tract (LVOT), leaflets, interleaflet triangles, and ascending aorta.

In order to obtain these regions, the deformation anatomy (after simulated device deployment) was firstly divided into the anatomical sections mentioned above. Then, each element of the simulated skirt was attributed to one of these anatomical regions and the distance between the skirt and the anatomy was calculated. This was done by searching the anatomy element in the normal direction to each skirt element. Apposition and malapposition were then attributed to each element based on the distance (apposed if distance smaller than 1mm, malapposed otherwise). Finally, the skirt was projected in 2D and the apposed and malapposed areas computed for each anatomical section. A visual overview of the separation of the skirt into sections (both anatomical and apposition) is shown in Figure 2. The obtained area values were grouped according to the aortic valve morphology: tricuspid (TAV), bicuspid (all types, BAV), bicuspid type 0 (BAV0) and BAV type 1 (BAV1) using the Sievers classification [15].

**Figure 2.**
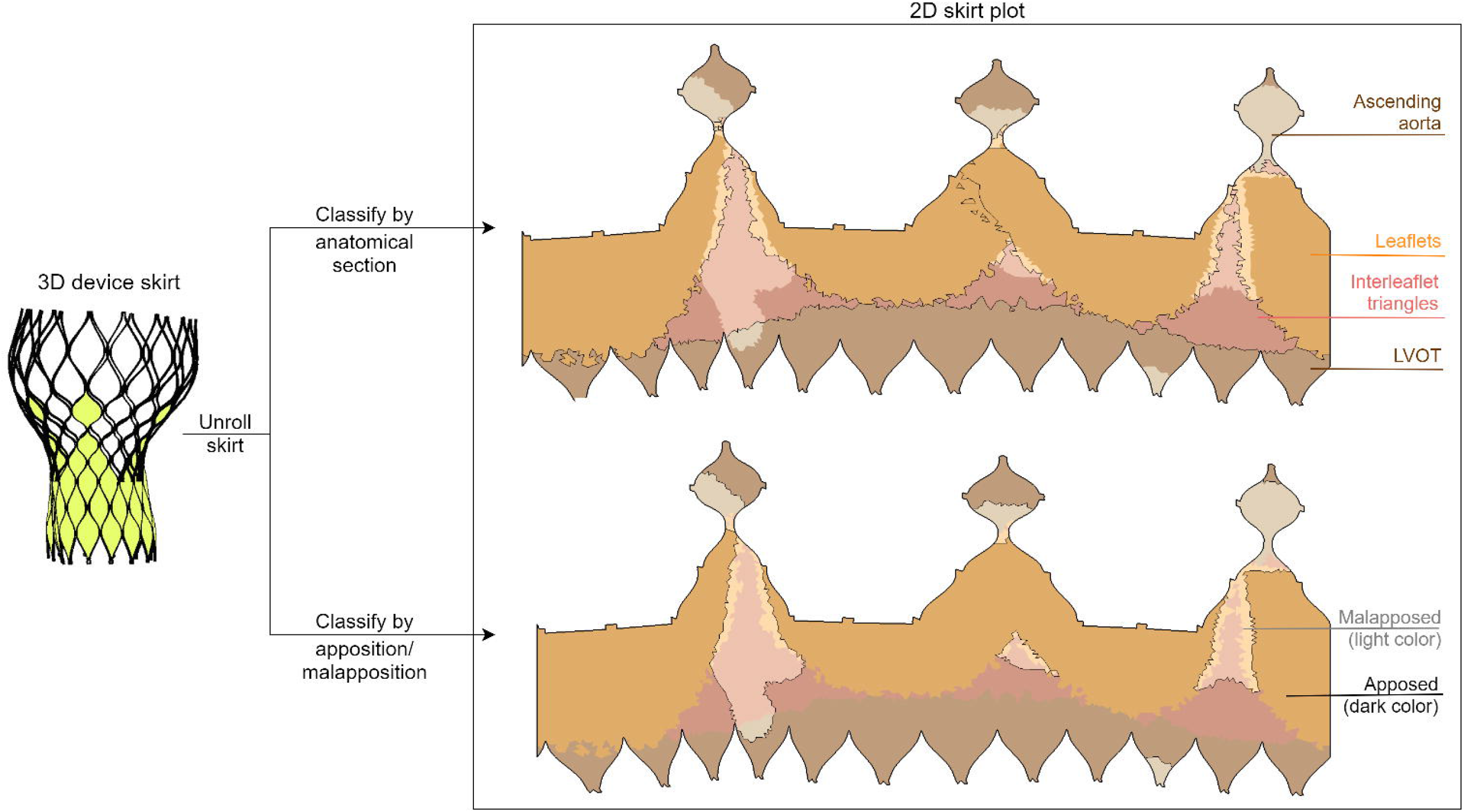
Illustration of the division of the two-dimensional deformed device skirt into anatomical (top panel) and apposition/malapposition (bottom panel) sections. This division is evidenced both with a border line and with a color palette, as depicted in the legend.

#### PVL comparison

Transthoracic Doppler echocardiography was used for the clinical PVL assessment. PVL was classified as none or trace, mild, or moderate based on the VARC-2 criteria. Observed PVL grades were compared to the predicted amount of skirt malapposition. The grades were also divided per valve morphology to detect possible patterns between the PVL severity and valve morphology.

## Statistical Analysis

Continuous variables are expressed as mean±SD. Correlation between predicted and observed continuous variables was analysed using the coefficient of determination (R^2^). Difference plots were constructed according to the Bland-Altman method. Comparisons within the sealing analysis were carried out using the paired Student t test or Mann–Whitney U test depending on the variable distribution. Baseline characteristics and anatomic parameters were analysed to explore the association with malapposition in interleaflet triangles. Only variables yielding a p value < 0.1 were included in the stepwise multivariate linear regression analysis. Statistical significance was defined as a two-tailed p < 0.05. Statistical analysis was performed with SciPy Stats, a Python module for probability functions and statistical distributions.

## Results

A total of 43 patients were included in the study. There was no significant difference in age between BAV and TAV patients (BAV: 76.4±7.1 years old vs. TAV: 79.4±6.2 years old, p = 0.164) or other baseline characteristics, Table 1. Of these 43 patients, 26 patients were BAV patients with 11 patients were type 0 and 15 were type 1. For the BAV patients, the sizing index (ratio of device size to perimeter derived diameter) was lower when compared with TAV patients (BAV: 1.04±0.09 vs. TAV: 1.11±0.07, p = 0.018).

**Table 1.** Patient Characteristics.

|  | **BAV type 0** | **BAV type 1** | **BAV** | **TAV** | **p Value** |
| --- | --- | --- | --- | --- | --- |
|  | **n = 11** | **n = 15** | **n = 26** | **n = 17** |  |
| Age (yrs) | 77.8±5.6 | 75.4±8.1 | 76.4±7.1 | 79.4±6.2 | 0.164 |
| Male | 5(45.5) | 12(80.0) | 17(65.4) | 9(52.9) | 0.528 |
| Height (cm) | 159.1±6.5 | 165.7±6.5 | 162.9±7.2 | 162.2±8.8 | 0.780 |
| Weight (kg) | 59.0±8.0 | 64.9±9.4 | 62.4±9.1 | 60.4±11.8 | 0.544 |
| Body Mass Index (kg/m<sup>2</sup>) | 23.31±3.01 | 23.59±2.94 | 23.47±2.91 | 22.85±3.59 | 0.542 |
| STS | 5.29±3.12 | 5.15±2.56 | 5.21±2.75 | 8.33±5.95 | 0.124* |
| Echocardiography |  |  |  |  |  |
| Left ventricular ejection fraction (%) | 54.9±14.9 | 55.0±12.7 | 55.0±13.3 | 54.8±17.3 | 0.593* |
| Aortic valve area (cm<sup>2</sup>) | 0.53±0.21 | 0.66±0.15 | 0.60±0.19 | 0.62±0.21 | 0.726 |
| Mean gradient (mmHg) | 59.4±19.4 | 56.5±15.2 | 57.7±16.8 | 52.9±11.5 | 0.478* |
| Max velocity (m/s) | 4.98±0.90 | 4.67±0.94 | 4.80±0.92 | 4.70±0.42 | 0.526* |
| Multi-slice computed tomography |  |  |  |  |  |
| Max annulus diameter (mm) | 27.4±3.5 | 29.6±3.0 | 28.7±3.4 | 27.4±3.2 | 0.240 |
| Min annulus diameter (mm) | 21.7±3.5 | 22.7±3.4 | 22.3±3.4 | 21.1±2.3 | 0.209 |
| Mean annulus diameter (mm) | 24.5±3.4 | 26.2±3.1 | 25.5±3.3 | 24.3±2.7 | 0.212 |
| Perimeter derived diameter (mm) | 24.7±3.3 | 26.5±3.1 | 25.8±3.3 | 24.7±3.0 | 0.312 |
| Area derived diameter (mm) | 24.3±3.3 | 26.0±3.1 | 25.3±3.2 | 24.2±2.9 | 0.285 |
| Calcium volume (mm<sup>3</sup>) | 1210.7±778.0 | 1213.5±676.6 | 1212.4±701.6 | 781.8±576.6 | 0.094 |
| Procedural characteristics |  |  |  |  |  |
| Implanted depth (mm) | 6.2±3.3 | 6.1±4.2 | 6.1±3.7 | 7.4±3.1 | 0.236 |
| Device size |  |  |  |  | 0.586 |
| 23mm | 3(27.3) | 1(6.7) | 4(15.4) | 1(5.9) |  |
| 26mm | 7(63.6) | 8(53.3) | 15(57.7) | 11(64.7) |  |
| 29mm | 0(0.0) | 5(33.3) | 5(19.2) | 2(11.8) |  |
| 32mm | 1(9.1) | 1(6.7) | 2(7.7) | 3(17.6) |  |
| Sizing index§ | 1.05±0.11 | 1.03±0.09 | 1.04±0.09 | 1.11±0.07 | 0.018 |
| Postprocedural outcomes |  |  |  |  |  |
| Mortality | 0(0.0) | 0(0.0) | 0(0.0) | 0(0.0) | - |
| Stroke | 0(0.0) | 0(0.0) | 0(0.0) | 0(0.0) | - |
| MI | 0(0.0) | 0(0.0) | 0(0.0) | 0(0.0) | - |
| PVL III/IV | 3(27.3) | 2(13.3) | 5(19.2) | 1(5.9) | 0.376 |
| Pacemaker Implantation | 1(9.1) | 1(6.7) | 2(7.7) | 2(11.8) | 1.000 |

§Sizing index = (device size) / (perimeter-based diameter).
\* Mann-Whitney U test was used.
Data are presented as no. (%) and mean ± SD.
BAV, bicuspid aortic valve; MI, myocardial infarction; PVL, paravalvular leakage; TAV, tricuspid aortic valve.

### Comparison of Observed and Predicted Parameters

The mean differences and coefficients of determination between the measurements extracted from the post-operative and simulated device are summarized in Table 2. This is presented for each type of measurement for all levels of the device combined. A high coefficient of determination was obtained for all measurements (≥ 0.90). All dimensions were slightly underestimated by the model, but the mean differences are negligible. Correlation and difference plots for each type of measurement are presented in Figure 3a and Figure 3b.

**Table 2.** Mean (±SD) difference between the measurements of the observed (post-operative) and the simulated (model) device and respective R-squared coefficient for the different levels of the device.

| Measurement | Mean Difference<br>(Post-op - Model) | R<sup>2</sup> |
| --- | --- | --- |
| Dmax (mm) | 0.10±1.42 | 0.90 |
| Dmin (mm) | 0.06±1.55 | 0.90 |
| Perimeter (mm) | 0.34±3.25 | 0.95 |
| Area (mm<sup>2</sup>) | 0.96±41.14 | 0.96 |

**Figure 3.**
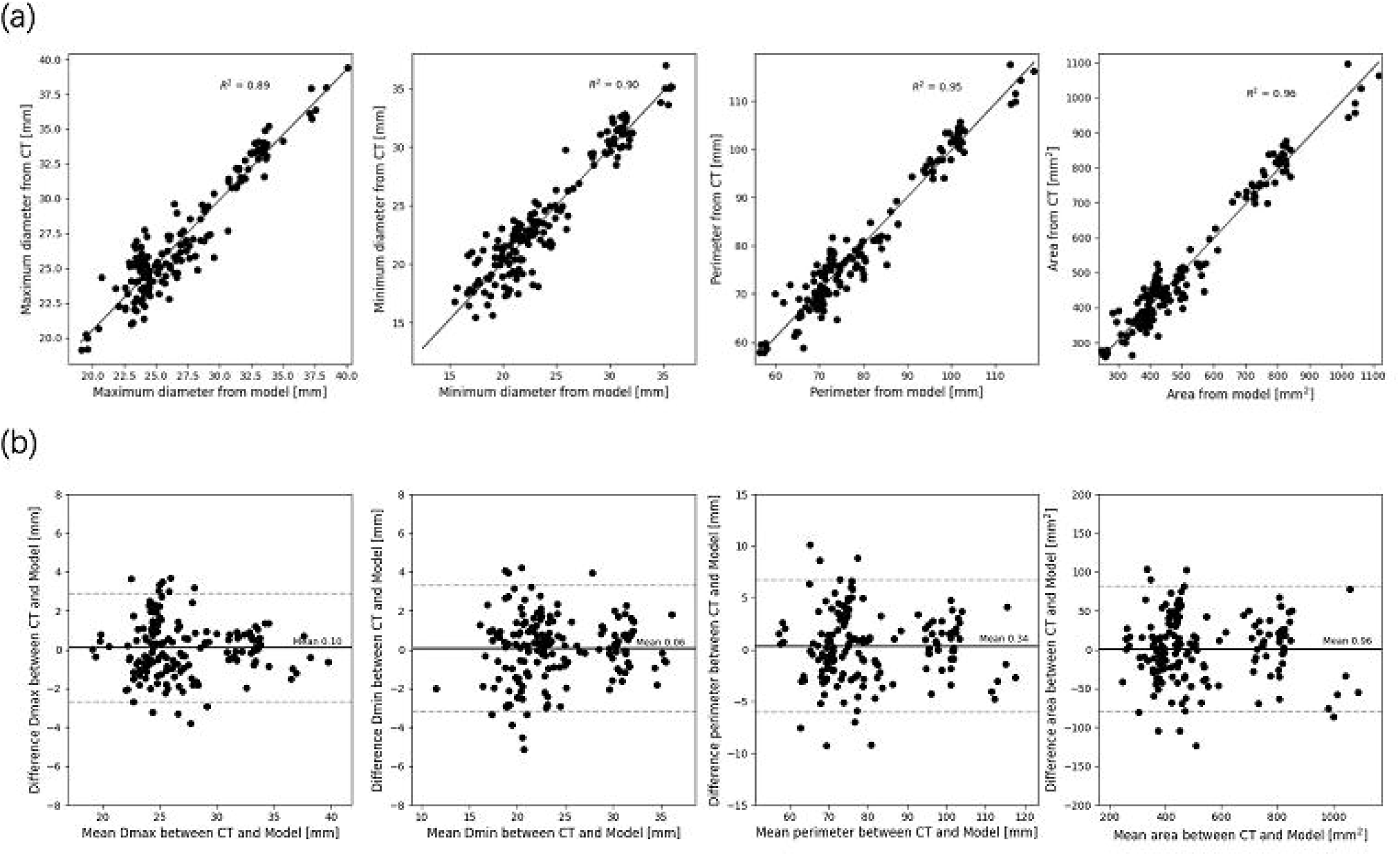
(a) Correlation and (b) difference plots for obtained device measurements vs predicted device measurements for all device levels.

Echocardiography showed none or trace post-operative PVL in 13 patients, mild PVL in 24 and moderate PVL in 6. Figure 4 shows a comparison of predicted skirt malapposition for patients with none to trace, mild and moderate PVL. The amount of skirt malapposition is higher for patients with a higher degree of clinically assessed PVL (Supplementary Table 1). A comparison of post-operative PVL assessment for patients with different valve morphologies is shown in Figure 5. Moderate PVL was more frequent for BAV cases (19.2% vs TAV 5.9%), with BAV0 having the highest incidence of moderate PVL (27.3% for BAV0 vs 13.3% for BAV1, respectively).

**Figure 4.**
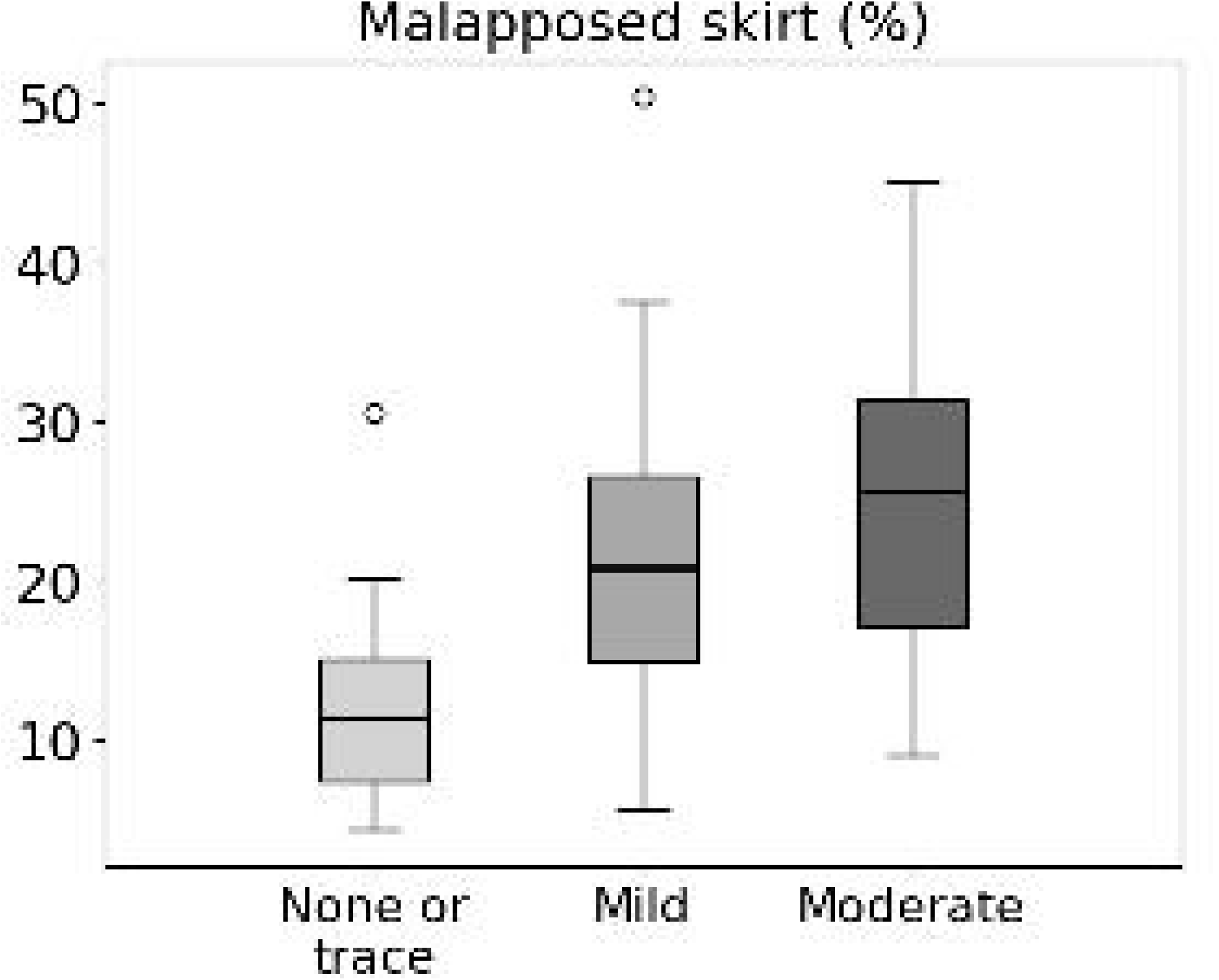
Box and whiskers diagram showing predicted skirt malapposition for patients with none or trace, mild and moderate PVL. Extreme values are presented as small circles (o).

**Figure 5.**
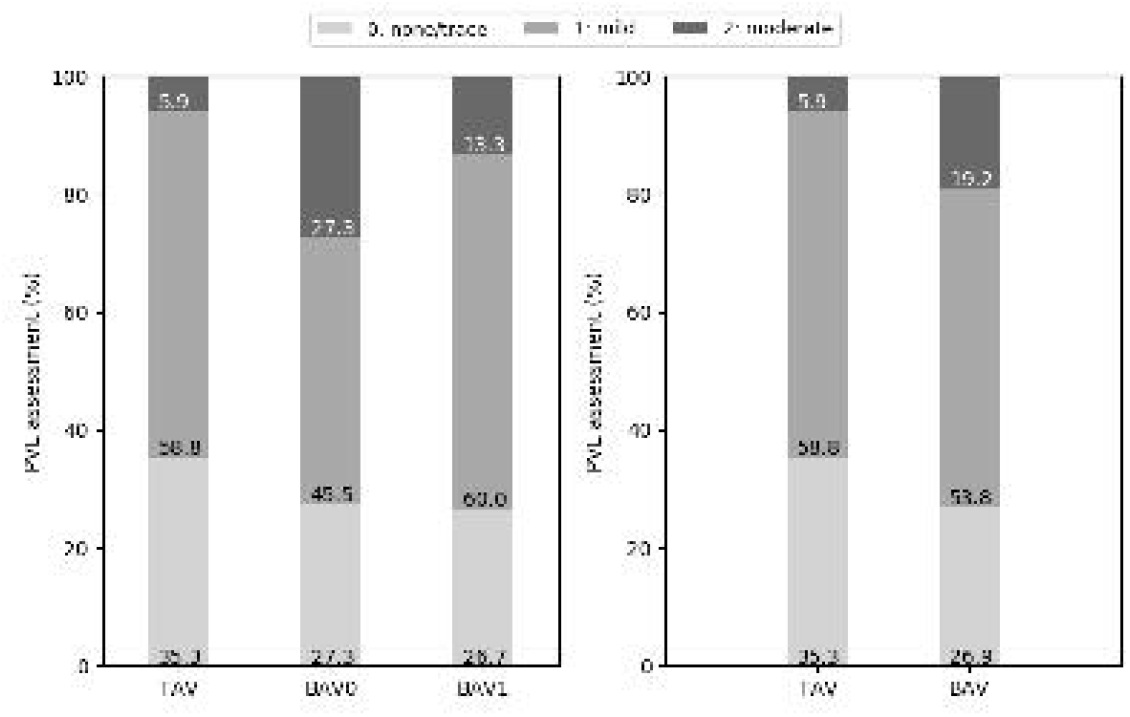
Post-operative PVL grading (none or trace, mild and moderate) for the different types of valve morphologies: TAV, BAV, BAV0 and BAV1.

### Comparison of sealing behaviour

A representative TAV (total skirt malapposition of 4.4%, no PVL) and BAV1 (total skirt malapposition of 20.9%, mild PVL) case are depicted in Figure 6. A cross-section of the pre-operative CT scan at the aortic annular plane and a 3D reconstruction illustrate the morphology of the valves. For each valve, the 2D skirt is also shown with the apposition borders highlighted, evidencing the larger area of malapposition in the BAV1 case (relatively to TAV). For the BAV1 case, PVL channels are visible in the interleaflet triangles region.

**Figure 6.**
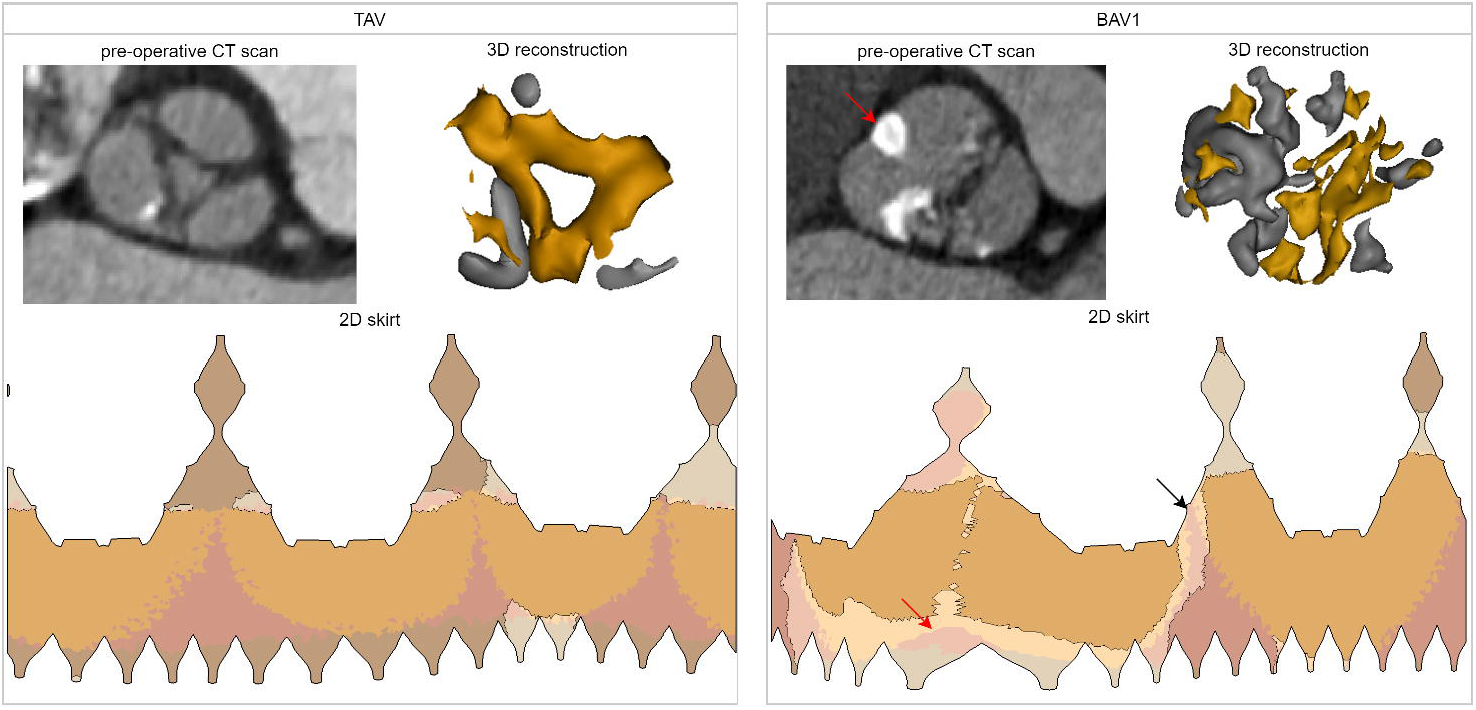
Analysis of two cases of the cohort with different valve morphologies: TAV (total skirt malapposition of 4.4%, PVL grade 0) and BAV1 (total skirt malapposition of 20.9%, PVL grade 1). For each case, the following is shown: cross-section at the aortic annular plane in the pre-operative CT, three-dimensional reconstruction of the valve (and its calcifications) and distribution of the apposition/malapposition sections of the two-dimensional deformed device skirt. For the BAV1 case, the red arrow indicates the raphe and the black arrow the largest PVL channel. CT = computed tomography; 3D = three-dimensional; 2D = two-dimensional

An overview of all sealing analysis data for each valve morphology is provided in Table 3. The mean percentage of total malapposed skirt obtained for each anatomical section (LVOT, interleaflet triangles and leaflets) relatively to the total skirt is illustrated as bar plots for the different valve morphologies in Figure 7a. In this analysis, the values obtained for the ascending aorta section were not considered to simplify the analysis. More malapposition was obtained for BAV cases when compared to TAV cases (22.7 vs 15.5%, p < 0.05), and this is also true when comparing TAV cases to BAV type 0 and BAV type 1 cases separately. This seems mainly driven by a higher degree of malapposition in the interleaflet triangles section: 5.8% and 11.1% for TAV and BAV (p < 0.05), respectively.

**Table 3.** Overview of sealing analysis data for the different valve morphologies.

|  | BAV type 0<br>n = 11 | BAV type 1<br>n = 15 | BAV<br>n = 26 | TAV<br>n = 17 | p Value |
| --- | --- | --- | --- | --- | --- |
| LVOT |  |  |  |  |  |
| Apposed area (mm<sup>2</sup>) | 72.4±68.4 | 133.1±190.4 | 107.4±152.0 | 157.4±135.7 | 0.106* |
| Malapposed area (mm<sup>2</sup>) | 47.8±87.0 | 81.6±126.4 | 67.3±110.7 | 52.0±79.9 | 0.980* |
| Interleaflet triangles |  |  |  |  |  |
| Apposed area (mm<sup>2</sup>) | 146.4±95.2 | 123.8±75.5 | 133.4±83.4 | 175.1±73.4 | 0.100 |
| Malapposed area (mm<sup>2</sup>) | 146.1±60.1 | 100.5±45.7 | 119.8±56.1 | 65.5±36.4 | **0.001** |
| Leaflets |  |  |  |  |  |
| Apposed area (mm<sup>2</sup>) | 590.6±84.2 | 628.3±191.8 | 612.4±154.3 | 611.5±193.6 | 0.987 |
| Malapposed area (mm<sup>2</sup>) | 69.1±52.0 | 73.8±30.6 | 71.8±40.1 | 56.8±30.1 | 0.195 |
| Malapposition in total skirt (%) | 24.1±10.6 | 21.6±10.5 | 22.7±10.5 | 15.5±9.8 | **0.030** |
| Malapposition in LVOT (%) | 3.6±5.4 | 6.0±8.2 | 4.9±7.1 | 4.6±7.1 | 0.960* |
| Malapposition in Interleaflet triangles (%) | 14.0±6.4 | 9.0±4.0 | 11.1±5.7 | 5.8±2.8 | **0.001\*** |
| Malapposition in Leaflet (%) | 6.5±4.9 | 6.7±3.1 | 6.6±3.8 | 5.2±2.9 | 0.200 |
| Apposition in total skirt (%) | 75.9±10.6 | 78.4±10.6 | 77.3±10.5 | 84.5±9.8 | **0.030** |
| Apposition contribution LVOT (%) | 8.4±7.9 | 14.2±19.8 | 11.7±15.9 | 17.0±16.0 | 0.124* |
| Apposition contribution Interleaflet triangles (%) | 16.8±8.7 | 13.1±7.0 | 14.7±7.9 | 18.0±4.9 | 0.172* |
| Apposition contribution Leaflet (%) | 74.8±14.0 | 72.7±20.0 | 73.6±17.4 | 64.9±17.6 | 0.120 |

\* Mann-Whitney U test was used.
Data are presented as mean ± SD.
BAV, bicuspid aortic valve; LVOT, left ventricular outflow tract; TAV, tricuspid aortic valve.

**Figure 7.**
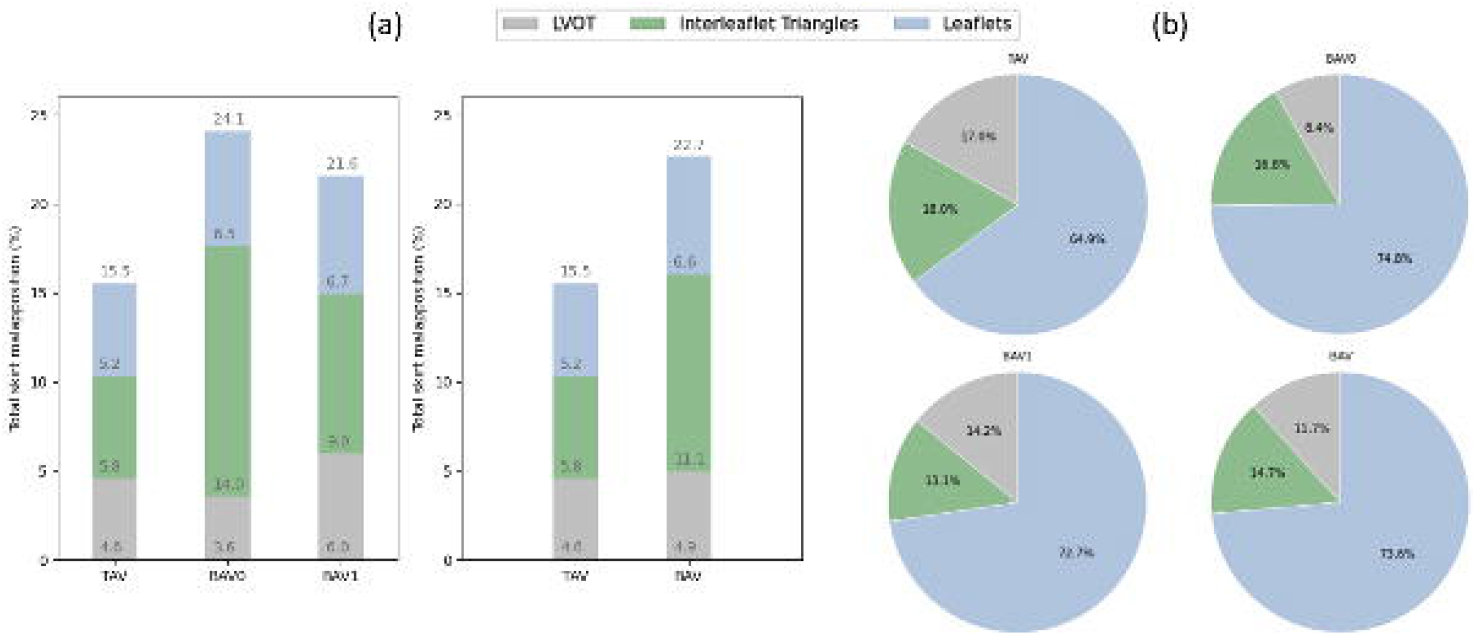
(a) Contribution of each anatomical section to the malapposition (mean, %) of the total skirt. (b) Contribution of each anatomical section to the apposition (mean, %) for the apposed section of the skirt. All these values are presented for the different types of valve morphologies: TAV, BAV, BAV0 and BAV1. TAV = tricuspid aortic valve; BAV = bicuspid aortic valve; BAV0 = type 0 bicuspid aortic valve; BAV1 = type 1 bicuspid aortic valve

The percentage of apposed skirt obtained out of the apposed section of the total skirt is illustrated as pie charts for each anatomical section and valve morphology in Figure 7b. There is a trend for a higher contribution of the left ventricular outflow tract (LVOT) to the apposition in TAV (17.0%) as compared to BAV cases (11.7%), and this difference is most pronounced for BAV type 0 patients (8.4%). In contrast, the leaflets seem to contribute more to apposition in BAV (73.6%) than in TAV cases (64.8%), and this is also true when looking at BAV type 0 and type 1 separately. However, no statistical significance was observed for these comparisons.

The multivariate linear regression analysis identified that BAV (p = 0.009) and sizing index (p = 0.034) as two independent predictors of malapposition in interleaflet triangles. The results of univariate and multivariate linear regression for association with malapposition in interleaflet triangles are presented in Supplementary Table 2.

## Discussion

In this study, we evaluated a patient-specific computer model of TAVR in Chinese BAV and TAV patients with the self-expanding Venus A-Valve using FEops HEARTguide (FEops, Ghent, Belgium). We compared the predicted THV frame deformation with postprocedural CT and found excellent correlation. Moreover, we observed a good agreement between predicted THV skirt malapposition and postoperative PVL based on echocardiography. Finally, we conducted a detailed sealing analysis and found that more malapposition was obtained in BAV patients when compared to TAV patients which was mainly driven by more malapposition at the location of the interleaflet triangles. Interestingly, the leaflets seem to be the main contributor to device sealing (apposition) not only in BAV but also in TAV cases.

### Validation of the modeling

Patient-specific computer simulation of TAVR has been previously described and validated, not only in TAV patients but also in BAV cases (FEops, Ghent, Belgium) [8–12]. These previous studies showed that computer simulation can accurately predict the THV frame deformation, severity of PVL, and potential occurrence of conduction abnormalities. However, these studies primarily focused on the Medtronic self-expanding and the Boston Scientific mechanically expandable THVs, and were all conducted by European hospitals. In this study, we employed the patient-specific computer simulation for the first time in a Chinese patient population with the self-expanding Venus A-Valve. Despite the higher radial force of the Venus A-Valve and the high calcium burden in this Chinese population, an excellent agreement between the predicted and observed dimensions of the valve frame was obtained [5,16]. Moreover, we compared predicted THV skirt malapposition and clinically assessed PVL, and found a good relationship.

These validated patient-specific computer simulations may help clinicians to better understand the risk of the procedure, and to optimize device sizing and positioning for each individual. This useful tool can also assist physicians recognizing patients who would benefit from TAVR and other patients for whom SAVR may be the preferred treatment. TAVR in mainland China is rapidly evolving, and the most widely used commercial THV is currently the Venus A-Valve [17,18]. Therefore, the presented study may be an important step to bring this technology to the Chinese physicians.

### Sealing behavior in BAV and TAV patients

TAVR in BAV patients has proven to be safe and effective, but patients need to be selected carefully and a widely accepted THV sizing strategy is still lacking. One key challenge of BAV disease is the increased anatomical heterogeneity as compared to TAV disease. In addition, there are important ethnic differences. In European populations, BAV type 1 with L-R coronary cusp fusion is most common, while in Asian populations, an unexpected high prevalence of type 0 was found [19,20]. As the deformed device skirt mainly interacts with three different regions of the aortic root anatomy, LVOT, leaflets, and interleaflet triangles, we performed a detailed analysis of the sealing behavior in these anatomical regions in BAV and TAV patients.

In the presented study, we found more malapposition in BAV patients when compared to TAV patients which was mainly driven by more malapposition in the interleaflet triangles. It should be emphasized that the pathologic landmark of BAV is always an absent or underdeveloped interleaflet triangle: a dysmorphic, underdeveloped interleaflet triangle is usually accompanied by a raphe, while the type 0 BAV is a valve with complete absence of one interleaflet triangle [21]. Another factor is that TAVR in BAV might result in uneven bioprosthetic valve frame expansion after THV deployment, and the deformed device (skirt) may not touch the interleaflet triangle under the restricted stent-frame expansion [19,22–24]. This may explain the higher amount of malapposition that we observed in the interleaflet triangle region in type 0 and type 1 BAV cases compared with TAV.

Another finding is that the leaflets seem to be the main contributor to device sealing, not only in BAV but also in TAV cases. The importance of the interaction between the supra annular structure and THV has already been discussed in previous studies [7,14,25–27]. Our present study based on patient-specific computer simulation further clarifies the crucial contribution of the leaflets to the supra-annular sealing. Overall, these results confirm that an assessment of the supra-annular structure is important for the adequate planning of TAVR in BAV cases. Moreover, in our present study, we found a trend for a higher contribution of the LVOT to the apposition in TAV as compared to BAV cases, and this difference is most pronounced for BAV type 0 patients. This result may be partially explained by the depth of implantation. As described in the baseline characteristics, BAV type 0 patients had a tendency of higher implantation than TAV. In addition, for BAV type 0 patients, the fish mouth like shape of the valve may result in an under expansion of the THV frame in the annular and sub-annular (LVOT) region, and thus reduce the device-tissue interaction in this region.

### Malapposition and PVL

The presented sealing analysis based on computational modeling may reflect the risk of PVL after TAVR. As showed in Figure 4, a higher amount of malapposition section seems related to the echo-based PVL grading. In addition, we observed a higher prevalence of moderate echocardiographic identified PVL in the BAV group which might be explained by the observed difference in sealing behavior between BAV and TAV cases. Understanding the sealing behavior of TAVR in BAV and TAV patients could assist physicians to comprehensively assess the risk of PVL and evaluate the interaction between supra-annular structure and THV stent frame.

## Limitations

This study was a small and retrospective single-center study. Due to the low number of patients with more than moderate PVL, no formal statistical analysis was performed. As a retrospective study, the transthoracic echocardiography was used to assess the clinical PVL and the location of PVL can’t be evaluated for the limitation of imaging quality.

## Conclusions

Patient-specific computer simulation of TAVR can be used in Chinese patients with the self-expanding Venus A-Valve. Transcatheter aortic valve sealing behavior is different between BAV and TAV patients with more malapposition at the location of the interleaflet triangles section for BAV cases.

## Data Availability

Data will be accessed after reasonable request.

## Contributors

J-AW, X-BL, and J-QF design the study. X-BL, J-QF, Y-XH, Q-FZ, Y-CG, X-PL, H-JL, and J-BJ conducted the study and performed the examinations. PM, GR, VO, TD, and J-QF performed the patient-specific computer simulation and statistical analysis. J-QF and PM performed the statistical analysis and wrote the manuscript. J-AW, LS, and PM revised the manuscript. All authors have read and approved the final version of the manuscript.

## Funding

This research project was partially funded by Venus Medtech.

## Competing interests

None declared.

## Patient and public involvement

Patients and/or the public were not involved in the design, conduct, reporting or dissemination plans of this research.

## Relationship with Industry

GR, TD and VO are employees of FEops NV. PM is co-founder of FEops NV. All other authors declare that there is no conflict of interest relevant to the submitted work.

## Nonstandard Abbreviations and Acronyms

AS: aortic stenosis
BAV: bicuspid aortic valve
CAD: computer aided design
CT: computed tomography
FEA: finite element analysis
HU: Hounsfield unit
DSCT: dual source computed tomography
TAV: tricuspid aortic valve
TAVR: transcatheter aortic valve replacement
TTE: transthoracic echocardiography

## Notes

### Competing Interest Statement

The authors have declared no competing interest.

### Author Declarations

The study was approved by the medical ethics committee of Second Affiliated Hospital of Zhejiang University and carried out according to the principles of the Declaration of Helsinki. All patients provided written informed consent for TAVR and the use of anonymous clinical, procedural, and follow-up data for research.

