## Supplementary Table for "Sealing Behaviour in Transcatheter Bicuspid and Tricuspid Aortic Valves Replacement through Patient-Specific Computational Modeling"

### Supplementary Tables

**Supplementary Table 1. Overview of sealing analysis data according to the severity of observed paravalvular leakage.**

| | Total<br>n = 43 | None or trace PVL<br>n = 13 | Mild PVL<br>n = 24 | Moderate PVL<br>n = 6 | p Value |
|---|---|---|---|---|---|
| LVOT | | | | | |
| Apposed area ($mm^2$) | 127.2±146.2 | 112.4±81.9 | 158.5±178.3 | 33.8±39.6 | 0.069* |
| Malapposed area ($mm^2$) | 61.2±98.9 | 19.5±27.6 | 89.3±122.0 | 39.2±54.5 | 0.104* |
| Interleaflet triangles | | | | | |
| Apposed area ($mm^2$) | 149.9±81.3 | 148.1±63.1 | 162.6±86.1 | 102.8±92.1 | 0.277 |
| Malapposed area ($mm^2$) | 98.4±55.7 | 66.3±44.3 | 109.1±46.7 | 124.9±84.7 | **0.033** |
| Leaflets | | | | | |
| Apposed area ($mm^2$) | 612.0±168.7 | 598.7±114.1 | 619.8±209.0 | 609.7±78.1 | 0.938 |
| Malapposed area ($mm^2$) | 65.9±36.9 | 34.5±25.6 | 76.8±29.4 | 90.0±45.1 | **0.000** |
| Malapposition in total skirt (%) | 19.8±10.7 | 12.7±7.3 | 22.3±10.1 | 25.5±12.8 | **0.009** |
| Malapposition in LVOT (%) | 4.8±7.0 | 1.9±2.6 | 6.7±8.5 | 3.7±5.0 | 0.146* |
| Malapposition in Interleaflet triangles (%) | 9.0±5.4 | 7.1±5.4 | 9.2±4.0 | 12.6±8.8 | 0.135* |
| Malapposition in Leaflet (%) | 6.0±3.5 | 3.7±3.1 | 6.4±2.4 | 9.3±5.2 | **0.002** |
| Apposition in total skirt (%) | 80.2±10.7 | 87.3±7.3 | 77.7±10.1 | 74.5±12.8 | **0.009** |

| | | | | | |
|---|---|---|---|---|---|
| Apposition in LVOT (%) | 13.8±16.0 | 12.8±9.9 | 16.9±19.3 | 4.0±4.3 | 0.080* |
| Apposition in Interleaflet triangles (%) | 16.0±7.0 | 16.6±6.3 | 16.6±7.0 | 12.5±8.6 | 0.288* |
| Apposition in Leaflet (%) | 70.2±17.8 | 70.7±13.8 | 66.6±19.7 | 83.5±12.0 | 0.113 |

* Kruskal Wallis test was used.

Data are presented as mean ± SD.

LVOT, left ventricular outflow tract; PVL, paravalvular leakage.

**Supplementary Table 2. Association of the Relevant Variables with Malapposition in Interleaflet triangles**

| Variables | Univariate linear regression | | | Multivariate linear regression* | | |
|---|---|---|---|---|---|---|
| | Coefficients | Standard error | p Value | Coefficients | Standard error | p Value |
| BAV vs. TAV (ref.) | 5.346 | 1.481 | 0.001 | 4.144 | 1.518 | 0.009 |
| Implanted depth | -0.240 | 0.235 | 0.314 | | | |
| Sizing index | -26.286 | 8.289 | 0.003 | -18.174 | 8.258 | 0.034 |
| Calcium volume (mm3) | 0.003 | 0.001 | 0.009 | | | |
| STJ diameter | 0.506 | 0.197 | 0.014 | | | |

*Stepwise selection was used to do the multivariable linear regression

BAV, bicuspid aortic valve; SOV, sinus of Valsalva; STJ, sino-tubular Junction; TAV, tricuspid aortic valve.
